# Evaluation of the concentration and profile of serum proteins and immunoglobulins in Mexican patients with advanced gastric cancer

**DOI:** 10.1101/19011759

**Authors:** María Alicia Díaz y Orea, Héctor Adrián Díaz Hernández, Rogelio Gonzalez Lopez, Eduardo Gómez Conde, Maria Elena Cárdenas Perea, Mónica Heredia Montaño, María José de los Ángeles Tapia Roldán, Emmanuel Marcelino Juarez Alvarado

## Abstract

**Introduction:** Gastric cancer remains an important health problem. It’s molecular mechanisms and interactions with the immune system are still not well elucidated. Therefore, we aimed to evaluate the concentration and profile of serum proteins and immunoglobulins in Mexican patients with advanced gastric cancer.

**Materials and methods:** We performed a descriptive study. Adult patients from both sexes were included. The problem group was formed of patients with advanced gastric cancer and the control group was formed of healthy subjects. Demographic data, gastric cancer histological type and stage of tumor node metastasis (TNM) system were recorded. The profile and concentration of serum proteins and immunoglobulins was determined and analyzed in different subgroups classified by sex, histologic type and stage of TNM system. To compare the concentrations of serum proteins and immunoglobulins the ANOVA test was performed. A p <0.05 was considered statistically significant.

**Results:** We included 88 patients with advanced gastric cancer and 74 healthy controls. There were no differences in demographic data among the groups, and the most common gastric cancer type was the diffuse (67.04%). Women with gastric cancer from any type presented higher levels of immunoglobulin G (IgG) compared with the control group (p <0.001) and men with gastric cancer of intestinal type in TNM stage III presented higher levels of IgG compared with it’s counterpart of diffuse type (p <0.001). Also, patients with intestinal type gastric cancer presented higher concentrations of alpha-1 globulins compared with patients with the diffuse type (p <0.05). Finally, patients with diffuse gastric cancer TNM stage IV presented the lowest albumin/globulin ratios.

**Conclusion:** There is a greater concentration of serum IgG in some subgroups of patients with advanced gastric cancer, the concentration of alpha-1 globulins is different between the intestinal and diffuse types and the albumin/globulin ratio is lower in the diffuse type.

## Introduction

Gastric cancer (GC) remains an important health problem. It is one of the most common causes of cancer and one of the leading causes of cancer-related death worldwide [1]. The Lauren classification sub-classifies GC into diffuse, intestinal and mixed type, each one with different pathological characteristics and prognosis [2-5]. Despite the great advances that have been made in the diagnosis and treatment of gastric cancer, its prognosis remains poor as it is a disease with nonspecific clinical manifestations in early stages, and then, it is usually diagnosed in advanced stages when it is no longer possible to offer curative therapies [6]. Hence, it remains relevant to study the molecular mechanisms regulating the development of gastric cancer and to identify new molecular targets for early detection and targeted treatments.

Several experimental studies have been performed with this approach, the vast majority directed to the detection of over-expression or anomalies in surface molecules and molecules from cellular secretion. Many of the studies have not shown satisfactory results, however, many others have favored the development of new treatments [7]. The changes in the expression of the cell surface molecules of malignant cells leads to a loss in the immunological tolerance, which triggers immunological responses against them. Although these anomalous changed molecules can be identified both in the interior and in the surface of the malignant cells, the antigens on the surface of the tumor are the mains responsible of the tumor immunogenicity [8-9].

Some studies have been carried out on the evaluation of the interaction between gastric cancer and the immune response. The local humoral immunological response has been studied, and it has been observed that in the cytoplasm of the advanced gastric cancer tumor cells there is less immunoglobulin M (IgM) and immunoglobulin A (IgA) distribution than in the cells of early gastric cancer and intestinal metaplasia. Similarly, it has been observed that in the stroma of advanced gastric cancer there is less infiltration of positive plasma cells for immunoglobulin G, (IgG), IgM and IgA, than in the stroma of early gastric cancer [10]. However, tissue levels of IgA in the unaffected stomach of patients with gastric cancer are elevated compared to healthy controls [11]. Regarding the systemic humoral immunological response, it has been observed that patients with gastric cancer have lower levels of globulins and IgG in comparison with healthy patients [12]. Moreover, different correlations have been established between gastric cancer and the humoral immune response. For example, it has been observed that depending on the stage of the disease and the immunoglobulin allotypes, the levels of antibodies against the membrane glycoprotein mucin 1 (MUC1), which is overexpressed in gastric cancer tumor cells, may be elevated [13]. Also, high levels of antibodies against the Thomsen-Friedenreich glycotope related to MUC1 have been observed in some cases and associated with a less aggressive course of the disease [14].

To date, there is no study that has evaluated the levels of globulins and immunoglobulins in patients with advanced gastric cancer in relation to the Lauren’s classification, which would help in further understanding the humoral immune response in this disease. Therefore, we aimed to evaluate the concentration and profile of serum proteins and immunoglobulins in Mexican patients with intestinal and diffuse advanced gastric cancer.

## Materials and methods

### Study type and population

We performed an observational, transversal, comparative and analytical study. Patients of both sexes from 20 to 85 years were included. The problem group was formed of patients from the High Specialty Medical Unit “Manuel Ávila Camacho”, Puebla, Mexico, with the diagnosis of gastric cancer in stages III and IV, according to the tumor node metastasis (TNM) system. The control group was formed of healthy volunteers. We excluded patients with immunosuppressive states as diabetes mellitus, autoimmune diseases, primary or secondary immunodeficiencies and patients on chemotherapy or radiotherapy. The recruitment period was from August 1^st^, 2016 to July 31^st^, 2018.

### Ethics approval and informed consent

This study was approved by the research and ethics committee of the School of Medicine of the Meritorious Autonomous University of Puebla, Puebla, Mexico, whit number 526 and written informed consent was obtained from all participants.

### Quantification of total proteins

#### Total proteins

The concentration of total proteins was determined by spectrophotometry by the Biuret technique following the instructions of Krueziger [15,16]. The biuret method is based on the formation of a colored copper protein complex which is measured by spectroscopy absorbance at 540 nm. A standard concentration curve was made using bovine serum albumin (BSA) before each determination to compare the absorbances of the samples. The absorbance of the color produced was read at 540 nm. The total protein concentration was recorded in g/dL.

### Determination and quantification of protein fractions

#### Serum protein electrophoresis in cellulose acetate

The protein fractions were determined by electrophoresis in cellulose acetate according to the standard method of the Analysis of Serum Protein, with Veronal buffer, pH 8.6, at an ionic strength of 0.05 [17]. Staining was done with Ponceau red. The concentration of each fraction (albumin, alpha-1 globulin, alpha-2 globulin, beta globulin and gamma globulin) was determined in a Bio-Rad Gel Doc^™^ XR^+^ Gel Documentation System and reported in g/dL.

### Determination and quantification of immunoglobulins

#### Enzyme-Linked Immuno-Sorbent Assay (ELISA)

The concentrations of serum immunoglobulins were determined by the Enzyme-linked Immunosorbent Assay (ELISA). The ELISA kits of eBioscience™ for Human IgM, IgG and IgA total (ELISA Ready-SET-Go!™ Kit) were used following the provider’s instructions. The concentration of the IgM, IgG and IgA values were recorded in mg/dL.

### Statistical analysis

We determined a sample size of 88 patients to achieve an alpha error of 0.05 and a power of 90%. We recorded general variables of age, sex, gastric cancer type of Lauren’s classification and stage of TNM classification. Demographic data are presented as numbers with percentages for categorical variables and mean with standard deviation for numerical variables. For the comparison between the groups, the X^2^ test and Student’s-t test were used as appropriate. The values of the determinations of total protein, albumin, alpha-1 globulin, alpha-2 globulin, beta globulin, gamma globulin, IgM, IgG and IgA were recorded. For the comparison between groups of the values of proteins, its fractions and immunoglobulins, the ANOVA test for three groups was performed. A p <0.05 was considered as statistically significant. The SPSS software was used.

## Results

We included 88 patients with gastric cancer and 74 healthy controls. Among the patients with gastric cancer, 24 (27.27%) were intestinal type, 59 (67.04%) were diffuse type and 5 (5.68%) were not classified. There were no differences between age and sex among the different gastric cancer types; however, there were more patients with stage IV disease in the intestinal type group (Table 1).

**Table 1.**
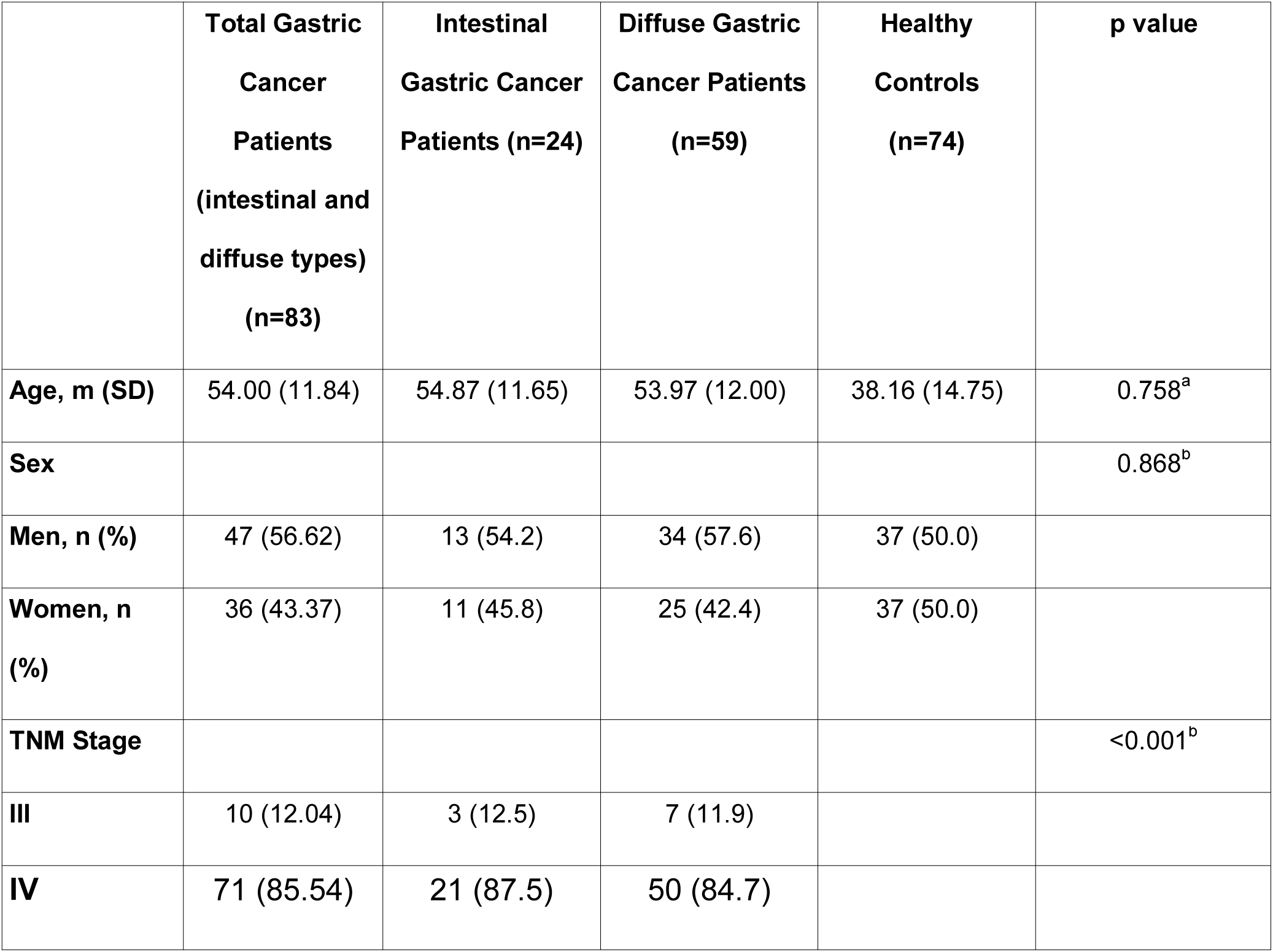
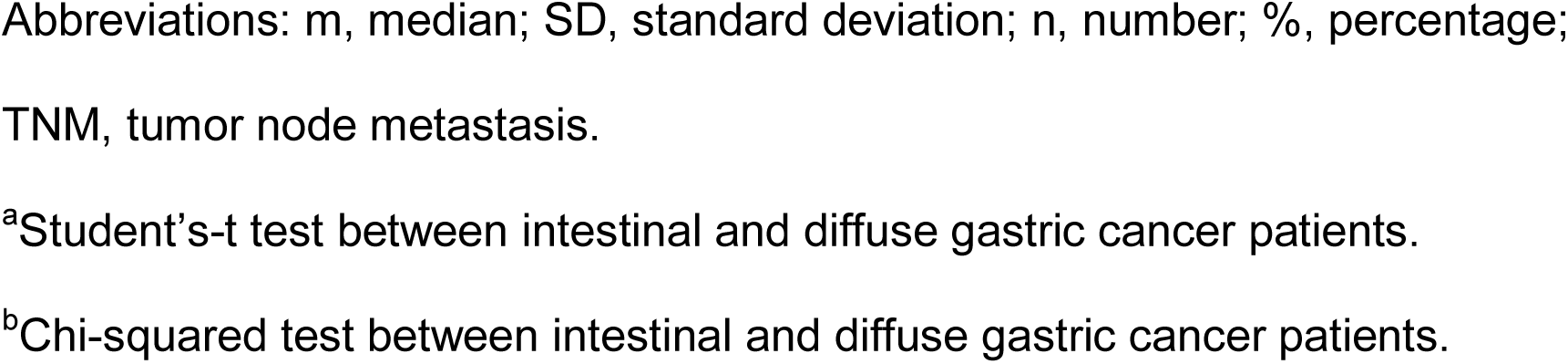
Demographic characteristics of patients with gastric cancer from the different types.

|  | **Total Gastric Cancer Patients (intestinal and diffuse types) (n=83)** | **Intestinal Gastric Cancer Patients (n=24)** | **Diffuse Gastric Cancer Patients (n=59)** | **Healthy Controls (n=74)** | **p value** |
| --- | --- | --- | --- | --- | --- |
| **Age, m (SD)** | 54.00 (11.84) | 54.87 (11.65) | 53.97 (12.00) | 38.16 (14.75) | 0.758<sup>a</sup> |
| **Sex** |  |  |  |  | 0.868<sup>b</sup> |
| **Men, n (%)** | 47 (56.62) | 13 (54.2) | 34 (57.6) | 37 (50.0) |  |
| **Women, n (%)** | 36 (43.37) | 11 (45.8) | 25 (42.4) | 37 (50.0) |  |
| **TNM Stage** |  |  |  |  | <0.001<sup>b</sup> |
| **III** | 10 (12.04) | 3 (12.5) | 7 (11.9) |  |  |
| **IV** | 71 (85.54) | 21 (87.5) | 50 (84.7) |  |  |

Abbreviations: m, median; SD, standard deviation; n, number; %, percentage; TNM, tumor node metastasis.
<sup>a</sup>Student's-t test between intestinal and diffuse gastric cancer patients.
<sup>b</sup>Chi-squared test between intestinal and diffuse gastric cancer patients.

### Patients with gastric cancer compared with healthy controls

We compared the concentration of the serum proteins and immunoglobulins between the patients with gastric cancer and the healthy controls. Only the women with gastric cancer presented higher levels of IgG compared with the women healthy control group, the remainder presented no differences. All comparisons are shown in Table 2.

**Table 2.** Levels of proteins, albumin, globulins and immunoglobulins between patients with gastric cancer and healthy controls.

|  | Gastric Cancer Patients | Healthy controls | p value |
| --- | --- | --- | --- |
| **Total** | (n=88) | (n=74) |  |
| **Proteins (g/dL), m (SD)** | 6.62 (1.87) | 7.84 (0.94) | >0.05<sup>a</sup> |
| **Albumin (g/dL), m (SD)** | 3.64 (1.20) | 4.39 (0.66) | >0.05<sup>a</sup> |
| **Alpha-1 globulin (g/dL), m (SD)** | 0.18 (0.15) | 0.14 (0.10) | >0.05<sup>a</sup> |
| **Alpha-2 globulin (g/dL), m (SD)** | 0.53 (0.22) | 0.53 (0.16) | >0.05<sup>a</sup> |
| **Beta globulin (g/dL), m (SD)** | 0.54 (0.24) | 0.69 (0.21) | >0.05<sup>a</sup> |
| **Gamma globulin (g/dL), m (SD)** | 1.68 (0.88) | 1.84 (0.42) | >0.05<sup>a</sup> |
| **IgM (g/dL), m (SD)** | 121.35 (44.10) | 142.64 (42.58) | >0.05<sup>a</sup> |
| **IgG (g/dL), m (SD)** | 2102.41 (526.15) | 2065.77 (438.82) | >0.05<sup>a</sup> |
| **IgA (g/dL), m (SD)** | 356.27 (29.78) | 356.68 (25.22) | >0.05<sup>a</sup> |
| **Men** | (n=49) | (n=37) |  |
| **Proteins (g/dL), m (SD)** | 6.94 (2.10) | 8.11 (1.08) | >0.05<sup>a</sup> |
| **Albumin (g/dL), m (SD)** | 3.82 (1.34) | 4.51 (1.04) | >0.05<sup>a</sup> |
| **Alpha-1 globulin (g/dL), m (SD)** | 0.17 (0.14) | 0.44 (0.34) | >0.05<sup>a</sup> |
| **Alpha-2 globulin (g/dL), m (SD)** | 0.55 (0.24) | 0.88 (0.83) | >0.05<sup>a</sup> |
| **Beta globulin (g/dL), m (SD)** | 0.54 (0.25) | 1.02 (0.21) | >0.05<sup>a</sup> |
| **Gamma globulin (g/dL), m (SD)** | 1.80 (0.97) | 1.24 (0.30) | >0.05<sup>a</sup> |
| **IgM (g/dL), m (SD)** | 117.79 (42.77) | 132.29 (47.93) | >0.05<sup>a</sup> |
| **IgG (g/dL), m (SD)** | 2016.04 (546.81) | 2121.35 (446.26) | >0.05<sup>a</sup> |
| **IgA (g/dL), m (SD)** | 361.20 (31.34) | 360.99 (22.16) | >0.05<sup>a</sup> |
| **Women** | (n=39) | (n=37) |  |
| **Proteins (g/dL), m (SD)** | 6.32 (1.49) | 7.56 (0.69) | >0.05<sup>a</sup> |
| **Albumin (g/dL), m (SD)** | 3.42 (0.96) | 4.06 (0.84) | >0.05<sup>a</sup> |
| **Alpha-1 globulin (g/dL), m (SD)** | 0.18 (0.15) | 0.37 (0.18) | >0.05<sup>a</sup> |
| **Alpha-2 globulin (g/dL), m (SD)** | 0.51 (0.19) | 0.84 (0.23) | >0.05<sup>a</sup> |
| **Beta globulin (g/dL), m (SD)** | 0.53 (0.21) | 1.02 (0.23) | >0.05<sup>a</sup> |
| **Gamma globulin (g/dL), m (SD)** | 1.54 (0.73) | 1.26 (0.21) | >0.05<sup>a</sup> |
| **IgM (g/dL), m (SD)** | 125.74 (45.86) | 153.00 (34.05) | >0.05<sup>a</sup> |
| **IgG (g/dL), m (SD)** | 2208.71 (485.48) | 2030.00 (432.47) | <0.001<sup>a</sup> |
| **IgA (g/dL), m (SD)** | 350.20 (26.90) | 352.37 (27.58) | >0.05<sup>a</sup> |

Abbreviations: m, median; SD, standard deviation; n, number; %, percentage;
IgM, immunoglobulin M; IgG, immunoglobulin G; IgA, immunoglobulin A.
<sup>a</sup>One-way ANOVA.

### Patients with intestinal type gastric cancer compared with patients with diffuse type gastric cancer

We compared the concentration of the serum proteins and immunoglobulins between the patients with intestinal type gastric cancer and the patients with diffuse type gastric cancer. In the patients with intestinal type the concentrations of alpha-1 globulins were higher. The remainder groups of albumin, globulins and immunoglobulins did not present differences. All comparisons are shown in Table 3.

**Table 3.** Levels of proteins, albumin, globulins and immunoglobulins between patients with gastric cancer from the different types.

|  | **Intestinal Gastric**<br>**Cancer Patients** | **Diffuse Gastric**<br>**Cancer Patients** | **p value** |
| --- | --- | --- | --- |
| **Total** | (n=24) | (n=59) |  |
| **Proteins (g/dL), m (SD)** | 6.0 (1.46) | 7.01 (2.04) | <0.05<sup>a</sup> |
| **Albumin (g/dL), m (SD)** | 3.37 (1.02) | 3.79 (1.28) | >0.05<sup>a</sup> |
| Alpha-1 globulin (g/dL), m (SD) | 0.22 (0.19) | 0.15 (0.12) | <0.05<sup>a</sup> |
| Alpha-2 globulin (g/dL), m (SD) | 0.45 (0.10) | 0.55 (0.25) | >0.05<sup>a</sup> |
| Beta globulin (g/dL), m (SD) | 0.46 (0.20) | 0.56 (0.26) | >0.05<sup>a</sup> |
| Gamma globulin (g/dL), m (SD) | 1.48 (0.83) | 1.78 (0.93) | >0.05<sup>a</sup> |
| IgM (g/dL), m (SD) | 108.75 (33.36) | 122.67 (46.12) | >0.05<sup>a</sup> |
| IgG (g/dL), m (SD) | 2165.00 (537.12) | 2039.05 (527.56) | >0.05<sup>a</sup> |
| IgA (g/dL), m (SD) | 353.45 (31.49) | 359.22 (29.31) | >0.05<sup>a</sup> |

Abbreviations: m, median; SD, standard deviation; IgM, immunoglobulin M; IgG, immunoglobulin G; IgA, immunoglobulin A.
<sup>a</sup>One-way ANOVA.

### Patients with TNM stage III gastric cancer compared with patients with TNM stage IV gastric cancer

We compared the concentration of the serum proteins and immunoglobulins between the patients with TNM stage III gastric cancer and the patients with TNM stage IV gastric cancer. There were no differences in the levels of proteins, albumin, globulins and immunoglobulins between these groups. All comparisons are shown in Table 4.

**Table 4.** Levels of proteins, albumin, globulins and immunoglobulins between patients with gastric cancer from different stages.

|  | TNM Stage III | TNM Stage IV | p value |
| --- | --- | --- | --- |

| Total | (n=10) | (n=71) |  |
| --- | --- | --- | --- |
| **Proteins (g/dL), m (SD)** | 6.68 (1.64) | 6.62 (1.98) | >0.05<sup>a</sup> |
| **Albumin (g/dL), m (SD)** | 3.64 (0.89) | 3.65 (1.28) | >0.05<sup>a</sup> |
| **Alpha-1 globulin (g/dL), m (SD)** | 0.17 (0.14) | 0.17 (0.15) | >0.05<sup>a</sup> |
| **Alpha-2 globulin (g/dL), m (SD)** | 0.63 (0.19) | 0.51 (0.22) | >0.05<sup>a</sup> |
| **Beta globulin (g/dL), m (SD)** | 0.54 (0.25) | 0.53 (0.24) | >0.05<sup>a</sup> |
| **Gamma globulin (g/dL), m (SD)** | 1.55 (0.61) | 1.72 (0.93) | >0.05<sup>a</sup> |
| **IgM (g/dL), m (SD)** | 115.30 (30.86) | 120.91 (45.57) | >0.05<sup>a</sup> |
| **IgG (g/dL), m (SD)** | 2017.00 (648.57) | 2110.56 (515.76) | >0.05<sup>a</sup> |
| **IgA (g/dL), m (SD)** | 346.10 (36.58) | 357.22 (28.59) | >0.05<sup>a</sup> |

Five patients with undetermined gastric cancer type were excluded and 2 patients with gastric cancer TNM stage II were excluded. Abbreviations: TNM, tumor node metastasis; m, median; SD, standard deviation; IgM, immunoglobulin M; IgG, immunoglobulin G; IgA, immunoglobulin A.
<sup>a</sup>One-way ANOVA.

### Patients with intestinal type gastric cancer compared with patients with diffuse type gastric cancer, stratified by TNM stage

We performed an analysis to evaluate differences in the concentration of the serum proteins and immunoglobulins between the intestinal and diffuse types of gastric cancer, stratified by the TNM stage. Only, in the sub-group of men with gastric cancer from the intestinal type in TNM stage III there was an increased level of IgG. The remainder presented no differences. All comparisons are shown in Tables 5 and 6.

**Table 5.** Levels of proteins, albumin, globulins and immunoglobulins between patients with gastric cancer TNM stage III from the different types.

|  | **Intestinal Gastric**<br>**Cancer Patients** | **Diffuse Gastric**<br>**Cancer Patients** | **p value** |
| --- | --- | --- | --- |
| **Total** | (n=3) | (n=7) |  |
| **Proteins (g/dL), m (SD)** | 6.26 (2.31) | 6.85 (1.47) | >0.05<sup>a</sup> |
| **Albumin (g/dL), m (SD)** | 3.50 (1.20) | 3.70 (0.83) | >0.05<sup>a</sup> |
| **Alpha-1 globulin (g/dL), m (SD)** | 0.26 (0.14) | 0.14 (0.14) | >0.05<sup>a</sup> |
| **Alpha-2 globulin (g/dL), m (SD)** | 0.48 (0.07) | 0.69 (0.20) | >0.05<sup>a</sup> |
| **Beta globulin (g/dL), m (SD)** | 0.37 (0.03) | 0.62 (0.28) | >0.05<sup>a</sup> |
| **Gamma globulin (g/dL), m (SD)** | 1.59 (0.93) | 1.54 (0.51) | >0.05<sup>a</sup> |
| **IgM (g/dL), m (SD)** | 133.00 (49.12) | 107.71 (20.02) | >0.05<sup>a</sup> |
| **IgG (g/dL), m (SD)** | 2286.66 (641.27) | 1901.42 (664.79) | >0.05<sup>a</sup> |
| **IgA (g/dL), m (SD)** | 369.00 (19.05) | 336.28 (38.88) | >0.05<sup>a</sup> |
| **Men** | (n=2) | (n=4) |  |
| **Proteins (g/dL), m (SD)** | 7.35 (1.90) | 6.87 (1.63) | >0.05<sup>a</sup> |
| **Albumin (g/dL), m (SD)** | 4.04 (1.07) | 3.79 (0.95) | >0.05<sup>a</sup> |
| **Alpha-1 globulin (g/dL), m (SD)** | 0.34 (0.02) | 0.05 (0.08) | >0.05<sup>a</sup> |
| **Alpha-2 globulin (g/dL), m (SD)** | 0.49 (0.09) | 0.64 (0.15) | >0.05<sup>a</sup> |
| **Beta globulin (g/dL), m (SD)** | 0.39 (0.00) | 0.47 (0.13) | >0.05<sup>a</sup> |
| **Gamma globulin (g/dL), m (SD)** | 2.01 (0.82) | 1.68 (0.58) | >0.05<sup>a</sup> |
| **IgM (g/dL), m (SD)** | 109.50 (38.89) | 109.50 (25.52) | >0.05<sup>a</sup> |
| **IgG (g/dL), m (SD)** | 2630.00 (339.41) | 1552.50 (397.61) | <0.001<sup>a</sup> |
| **IgA (g/dL), m (SD)** | 374.50 (23.33) | 340.25 (49.50) | >0.05<sup>a</sup> |
| **Women** | (n=1) | (n=3) |  |
| **Proteins (g/dL), m (SD)** | 4.10 (0.00) | 6.83 (1.59) | ND |
| **Albumin (g/dL), m (SD)** | 2.42 (0.00) | 3.58 (0.83) | ND |
| **Alpha-1 globulin (g/dL), m (SD)** | 0.10 (0.00) | 0.25 (0.12) | ND |
| **Alpha-2 globulin (g/dL), m (SD)** | 0.46 (0.00) | 0.76 (0.27) | ND |
| **Beta globulin (g/dL), m (SD)** | 0.34 (0.00) | 0.81 (0.34) | ND |
| **Gamma globulin (g/dL), m (SD)** | 0.75 (0.00) | 1.35 (0.45) | ND |
| **IgM (g/dL), m (SD)** | 180.00 (0.00) | 105.33 (14.50) | ND |
| **IgG (g/dL), m (SD)** | 1600 (0.00) | 2366.66 (721.48) | ND |
| **IgA (g/dL), m (SD)** | 358.00 (0.00) | 331.00 (28.05) | ND |

In the sub-group of women the one-way ANOVA test could not be performed because there was 1 patient with intestinal type gastric cancer. Abbreviations: TNM, tumor node metastasis; m, median; SD, standard deviation; IgM, immunoglobulin M; IgG, immunoglobulin G; IgA, immunoglobulin A; ND, not determined.
<sup>a</sup>One-way ANOVA.

**Table 6.** Levels of proteins, albumin, globulins and immunoglobulins between patients with gastric cancer TNM stage IV from the different types.

|  | **Intestinal Gastric**<br>**Cancer Patients** | **Diffuse Gastric**<br>**Cancer Patients** | **p value** |
| --- | --- | --- | --- |
| **Total** | (n=21) | (n=50) |  |
| **Proteins (g/dL), m (SD)** | 5.97 (1.39) | 6.90 (2.13) | >0.05<sup>a</sup> |
| **Albumin (g/dL), m (SD)** | 3.36 (1.03) | 3.77 (1.37) | >0.05<sup>a</sup> |
| **Alpha-1 globulin (g/dL), m (SD)** | 0.22 (0.20) | 0.15 (0.12) | >0.05<sup>a</sup> |
| **Alpha-2 globulin (g/dL), m (SD)** | 0.44 (0.11) | 0.54 (0.25) | >0.05<sup>a</sup> |
| **Beta globulin (g/dL), m (SD)** | 0.47 (0.21) | 0.55 (0.25) | >0.05<sup>a</sup> |
| **Gamma globulin (g/dL), m (SD)** | 1.47 (0.84) | 1.82 (0.95) | >0.05<sup>a</sup> |
| **IgM (g/dL), m (SD)** | 105.28 (30.63) | 127.48 (49.34) | >0.05<sup>a</sup> |
| **IgG (g/dL), m (SD)** | 2147.61 (536.77) | 2095.00 (511.44) | >0.05<sup>a</sup> |
| **IgA (g/dL), m (SD)** | 351.23 (32.59) | 359.74 (26.69) | >0.05<sup>a</sup> |
| **Men** | (n=11) | (n=27) |  |
| **Proteins (g/dL), m (SD)** | 6.10 (1.59) | 7.29 (2.44) | >0.05<sup>a</sup> |
| **Albumin (g/dL), m (SD)** | 3.35 (1.08) | 4.01 (1.58) | >0.05<sup>a</sup> |
| **Alpha-1 globulin (g/dL), m (SD)** | 0.22 (0.12) | 0.16 (0.14) | >0.05<sup>a</sup> |
| **Alpha-2 globulin (g/dL), m (SD)** | 0.46 (0.11) | 0.57 (0.29) | >0.05<sup>a</sup> |
| **Beta globulin (g/dL), m (SD)** | 0.45 (0.25) | 0.57 (0.28) | >0.05<sup>a</sup> |
| **Gamma globulin (g/dL), m (SD)** | 1.58 (1.04) | 1.93 (1.06) | >0.05<sup>a</sup> |
| **IgM (g/dL), m (SD)** | 112.90 (26.27) | 118.51 (50.44) | >0.05<sup>a</sup> |
| **IgG (g/dL), m (SD)** | 2068.18 (363.53) | 2011.48 (601.04) | >0.05<sup>a</sup> |
| **IgA (g/dL), m (SD)** | 361.36 (29.21) | 361.22 (29.92) | >0.05<sup>a</sup> |
| **Women** | (n=10) | (n=23) |  |
| **Proteins (g/dL), m (SD)** | 5.83 (1.19) | 6.43 (1.63) | >0.05<sup>a</sup> |
| **Albumin (g/dL), m (SD)** | 3.36 (1.02) | 3.50 (1.02) | >0.05<sup>a</sup> |
| **Alpha-1 globulin (g/dL), m (SD)** | 0.22 (0.27) | 0.15 (0.08) | >0.05<sup>a</sup> |
| **Alpha-2 globulin (g/dL), m (SD)** | 0.43 (0.12) | 0.49 (0.19) | >0.05<sup>a</sup> |
| **Beta globulin (g/dL), m (SD)** | 0.50 (0.17) | 0.52 (0.21) | >0.05<sup>a</sup> |
| **Gamma globulin (g/dL), m (SD)** | 1.34 (0.57) | 1.71 (0.83) | >0.05<sup>a</sup> |
| **IgM (g/dL), m (SD)** | 96.90 (34.19) | 138.00 (46.92) | >0.05<sup>a</sup> |
| **IgG (g/dL), m (SD)** | 2235.00 (690.82) | 2193.04 (370.20) | >0.05<sup>a</sup> |
| **IgA (g/dL), m (SD)** | 340.10 (33.91) | 358.00 (22.87) | >0.05<sup>a</sup> |

Abbreviations: TNM, tumor node metastasis; m, median; SD, standard deviation;
IgM, immunoglobulin M; IgG, immunoglobulin G; IgA, immunoglobulin A.
<sup>a</sup>One-way ANOVA.

### The albumin/globulin ratio in the intestinal and diffuse types of gastric cancer, stratified by TNM stage

Finally, the proportion of albumin/globulin was evaluated among patients with advanced gastric cancer, stratified by the intestinal and diffuse types, and by the TNM stage. In this evaluation, patients with diffuse gastric cancer of TNM stage IV presented more frequently values ≤1.2 than patients with intestinal gastric cancer of TNM stage IV, which presented more frequently values ≥1.4, *p* ≤*0*.*5* (Dunn’s Multiple Comparisons Test). The analysis is shown in Table 7.

**Table 7.**
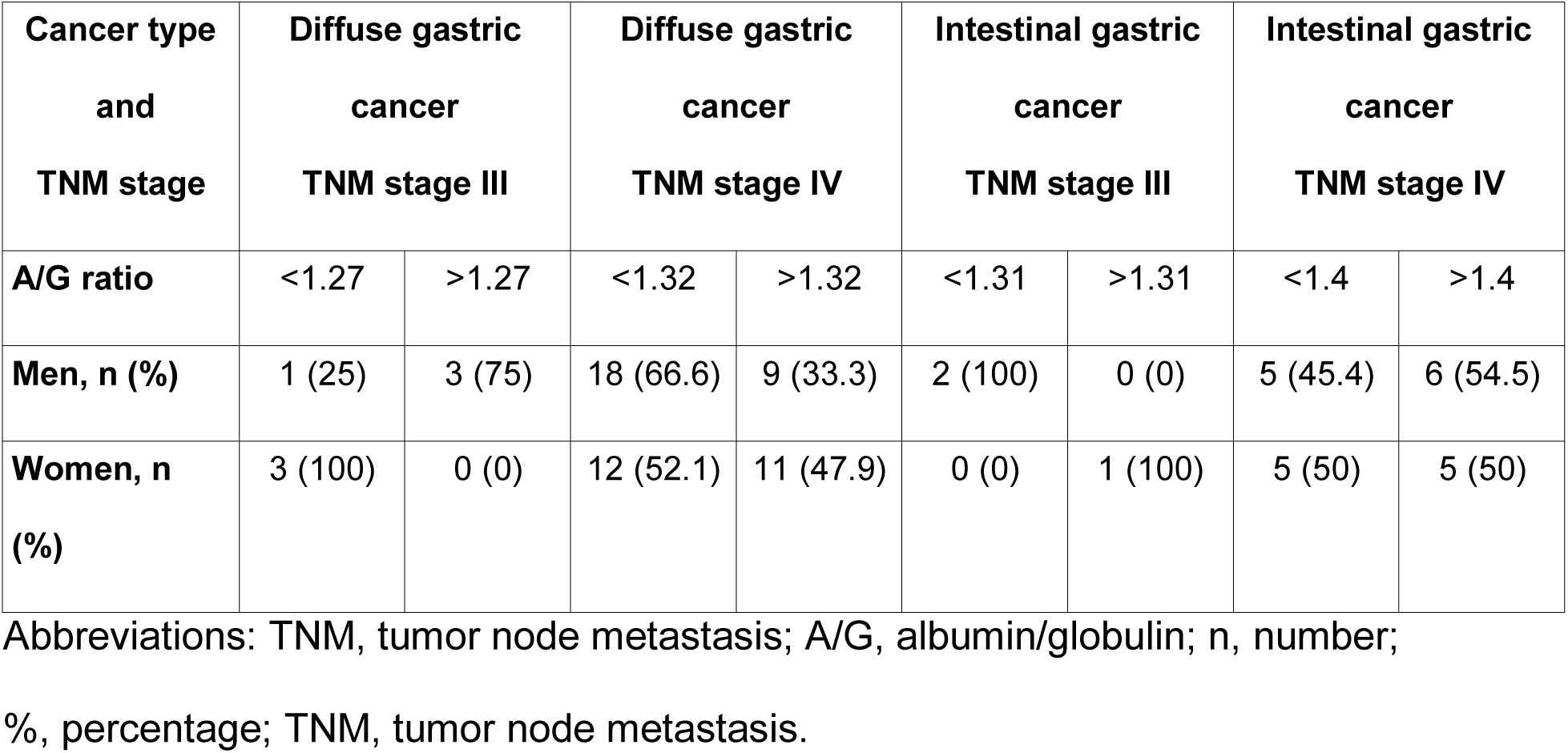
The albumin/globulin ratio of patients with gastric cancer TNM stages III and IV, from the different types.

## Discussion

In the demographic analysis, no relevant differences were observed between the groups of patients with intestinal and diffuse types of gastric cancer. The majority of patients with advanced gastric cancer were of the diffuse histological type (71%), which was expected as it is well described that gastric cancer of diffuse type has a more aggressive biological behavior, although lately, this aggressiveness has been attributed to a subgroup of diffuse gastric cancer that presents with “linitis plastica”, a particularly aggressive infiltrative pattern [18].

After comparing the concentrations of proteins and immunoglobulins between the groups of patients with advanced gastric cancer from different TNM stages (TNM stages III and IV), no differences were observed, and therefore, according to the concentration of serum proteins and immunoglobulins, both stages could be grouped into a single group of “advanced gastric cancer”. One of the most interesting findings from this study was the difference observed in IgG concentrations between healthy subjects and patients with advanced gastric cancer of certain subgroups. Compared with the control group, the subgroup of women with advanced gastric cancer (TNM stages III and IV) of any type (intestinal and diffuse), and the subgroup of men with advanced gastric cancer (TNM stage III) of the intestinal type, presented higher concentrations of serum IgG. These results suggest that in certain subgroups of patients with advanced gastric cancer there is an increase in the humoral immune response related to IgG. When comparing these results with previous studies that have evaluated other aspects of the humoral immune response in patients with gastric cancer, there are concordances and contradictions. On the one hand, in one study that evaluated the local humoral immune response, it was observed that in patients with advanced gastric cancer there was an increase in IgG and a depletion of IgM and IgA [10]. On the other hand, our results contradict the previously reported one in which it was observed that patients with gastric cancer had lower serum IgG levels than the control patients, however, this study included patients with gastric cancer of all stages, and in our study, we included Mexican patients with advanced gastric cancer (TNM stages III and IV), which may explain the difference, since in advanced gastric cancer there is a greater tumor burden and inflammation, which may explain an increased immunological response to antigens from the tumor cells or as consequence of the increased inflammatory response produced by the tumor, finally resulting in higher IgG serum levels in the patients with advanced gastric cancer [12]. Kurtenkov, et. al. have found elevated IgG anti-MUC1 autoantibodies in patients with gastric cancer and also found that these autoantibodies were associated with cancer progression [14]. We determined total IgG, which includes IgG antibodies and normal IgG, and found high values in patients with advanced gastric cancer, which could correlate with cancer progression too. This greater concentration of serum IgG, could also be an antibody against an antigen associated with the tumor that may favor the spread and metastasis, or may be an antibody produced by the tumor cells to promote its development.

Another interesting finding of this study was that when we compared the profile of serum proteins between patients with gastric cancer of different histologic types, an increase in the concentration of the alpha-1 globulin fraction was observed in the serum of patients with gastric cancer of the intestinal type. Therefore, further studies are required to evaluate the profile of the alpha-1 fraction of serum proteins in patients with gastric cancer of the intestinal type, with techniques that allow separation of all the fractions of proteins contained in the alpha-1 fraction such as polyacrylamide gel electrophoresis, ant then, determine if these are already known proteins that are elevated as acute phase reactants or if these are abnormal proteins secreted by the tumor cells. However, we found in 36.7% of women with diffuse gastric cancer stage IV and in 5.9% of women with diffuse gastric cancer stage III, low concentrations of alpha 1 globulin (α1 anti-trypsin), which may be due to a hereditary genetic predisposition in this type of cancer, and perhaps, because of the high percentage of patients in which alpha 1 globulin is found, diminished or absent, the Mexican population could carry a haplotype that makes them susceptible to this malignant disease. In breast cancer, α1 anti-trypsin, is a biomarker for early stages [19], in our patients it is decreased in advanced stages, so we could consider it as a biomarker of malignancy.

Further, we calculated the albumin/globulin ratio, this relationship is not only related to the nutritional status but to the inflammatory response [20]. The serum albumin is very important for the stabilization of DNA and inhibition of carcinogenesis. It is well established that elevated albumin levels are associated with a favorable prognosis of survival in several types of cancer [21]. Xue et al, related the pre-surgery values of the albumin/globulin ratio to the prognosis of gastric cancer, they found that a value <1.36 was associated with poor prognosis [22]. These values have further been related to sex, age and size of the tumor [23]. In our study, patients with diffuse gastric cancer stage IV predominated (74%), and in this patients we found that 66% had an albumin/globulin ratio of <1.2, hence, this relationship could be considered as a prognostic indicator of poor outcome from malignancy in Mexican population too.

Finally, in diffuse gastric cancer, the mutation of the gene of e-cadherin CH1, is considered and hereditary risk factor, and in our study population, we found a high prevalence of patients with diffuse gastric cancer in TNM stage IV, with low values of alpha 1 globulin (alpha-1 antitrypsin) and low albumin/globulin ratio, witch may be also related with a genetic predisposition. In some of these patients, we have made the determination of E-cadherin, and we have found it absent. For this reason, it’s possible that the Mexican population may have an allele, which makes them susceptible to diffuse gastric cancer type and an aggressive course of this disease, witch may be in relation to a polymorphism of the CDH1 gene specific to the Mexican population, as a genetic risk factor for diffuse gastric cancer type [24].

## Conclusion

In certain subgroups of patients with advanced gastric cancer there are higher serum concentrations of serum IgG, the concentration of serum alpha-1 globulins is different between the intestinal and diffuse types of gastric cancer and the albumin/globulin ratio is lower in the diffuse type of gastric caner. Further studies are needed to determine the meaning of these differences.

## Data Availability

All data is available in the records in the Laboratory of Experimental Immunology, Faculty of Medicine, Meritorious Autonomous University of Puebla, Puebla, Puebla, Mexico.

## Acknowledgments

This study was supported by the vice-rectory of research and postgraduate studies from the Meritorious Autonomous University of Puebla, Puebla, México. We thank all participating subjects for their cooperation in this study, and all the doctors who helped to recruit the subjects in our study.

